# Tracking viral RNA loads during in-sewer transport: insights from real-world field experiments

**DOI:** 10.64898/2026.02.09.26345892

**Authors:** Melissa Pitton, Charles Gan, Simon Bloem, David Dreifuss, Adrian Lison, Timothy R. Julian, Christoph Ort

## Abstract

Wastewater-based surveillance (WBS) is widely used to monitor respiratory viruses, yet uncertainties remain regarding how viral RNA concentrations in wastewater reflect infection dynamics. Specifically, diurnal variation in shedding and RNA losses during in-sewer transport can impact measured signals. We conducted a field study in a 5-km trunk sewer (travel time of one hour). Wastewater was sampled at the sewer inlet and outlet using autosamplers collecting time-proportional one-hour composite samples over 24 hours. The one-hour composite samples were analyzed for assessing intra-daily fluctuations, and 24-hour composites for signal change. Biofilms from the sewer-pipe walls were collected at three locations. Nucleic acids were extracted, and SARS-CoV-2, Influenza A/B, and Respiratory Syncytial Virus (RSV) RNA were quantified using a multiplex digital PCR assay. All viruses showed pronounced diurnal variation, with consistent morning load peaks. Viral RNA in the bulk liquid decreased during in-sewer transport, with modelled changes ranging from 15% to 72% across pathogens. Biofilms served as minor reservoirs of viral RNA; for SARS-CoV-2, sequencing revealed similarity between biofilm and bulk liquid RNA. Our study provides a full-scale assessment of in-sewer transport effects on viral RNA and highlights the need to account for complex in-sewer dynamics when interpreting WBS data.

## 1. Introduction

Wastewater-based surveillance (WBS) provides valuable insights into population-level disease dynamics, including those of infectious diseases caused by common respiratory viral pathogens, such as SARS-CoV-2, Influenza A and B viruses (IAV, IBV), and Respiratory Syncytial Virus (RSV). Typically, WBS relies on PCR-based molecular methods, such as qPCR and digital PCR (dPCR), to detect and quantify nucleic acids in wastewater as proxies for community disease incidence. Indeed, trends obtained from wastewater samples have been shown to strongly correlate with clinical case data and indicators, such as positivity rates^1–4^. Despite observed correlations between incidence and pathogen loads in wastewater, we are not yet able to infer incidence directly from pathogen loads due to several outstanding uncertainties^5,6^. These uncertainties arise at multiple stages of the WBS framework, spanning the five main conceptual steps: shedding dynamics, in-sewer fate and transport, sampling locations and strategies, sample processing, and molecular analyses.

In the context of shedding dynamics, infected individuals do not contribute equally to viral loads in wastewater. This variability arises from differences in factors such as the stage of the infection, the presence and severity of symptoms and the immune system^7–9^. Moreover, several shedding routes exist, including saliva, urine, and stools^10,11^.

Once excreted, viruses can undergo transformations in sewer networks and experience dilution before reaching a wastewater treatment plant (WWTP), attenuating the viral signal and leading to inherent variability of RNA loads at the sampling point. Viral stability can be influenced by the characteristics of the wastewater matrix, including factors like temperature and pH^12,13^. The heterogeneous and complex nature of wastewater must be considered, as viruses can exhibit different affinities for distinct compartments, such as the bulk liquid, suspended and settled solids, as well as biofilms^2,14–17^.

Sampling locations and strategies are factors to consider, as upstream sampling prior to dilution from surface runoff and industrial discharges can enhance sensitivity relative to downstream endpoints, such as WWTPs, where longer travel times may further increase RNA loss. Regarding sampling strategies, grab sampling provides a snapshot, which may not capture short-term fluctuations in viral concentrations or intra-day variability, and cannot determine these metrics with the desired or highest possible accuracy^18^. In contrast, 24-hour composite samples better represent temporal trends but are more susceptible to prolonged environmental exposure. Nonetheless, decisions on sampling location and strategy should take the study objective into account.

Following sample collection, wastewater samples are typically transported to laboratories, where storage conditions, particularly temperature and duration, play an important role in preserving viral integrity^19,20^. Viral RNA recovery is also highly influenced by the concentration and/or extraction methods used^21–23^, which could lead to substantial differences in the quantity of yielded nucleic acid.

Lastly, the sensitivity and specificity of the analytical techniques employed for RNA quantification, such as qPCR and dPCR, can introduce additional variability in the results^24^. This variability can arise from differences in assay efficiency^25^, the dynamic range of detection, and the susceptibility of each method to PCR inhibitors commonly present in wastewater matrices^26^.

Considering viral RNA dynamics is important for refining WBS and improving the accuracy of pathogen load estimations. Among the five main conceptual components of WBS, in-sewer transport remains one of the least understood, largely due to the complexity of studying full-scale sewer systems. Therefore, this study focused specifically on in-sewer transport, aiming to provide new insights through experiments conducted in a real-world sewer system. Using common respiratory viruses as targets, such as SARS-CoV-2, IAV, IBV, and RSV, we first examined diurnal variability in viral loads. We then evaluated the extent to which in-sewer transport affects viral levels by comparing measurements at two locations along a sewer. Finally, we investigated the influence of sewer biofilms on estimated loads. Our findings underscore the complexities of RNA dynamics and transport within sewer networks, emphasizing the need to account for these factors when interpreting WBS data for surveillance or for modelling applications.

## 2. Material and methods

### 2.1. Selected viral respiratory pathogens

This study focused on four specific respiratory viruses, severe acute respiratory syndrome coronavirus 2 (SARS-CoV-2), Influenza A Virus (IAV), Influenza B Virus (IBV), and Respiratory Syncytial Virus (RSV).

### 2.2. Sewer and subcatchment characteristics

All field experiments were conducted in a main trunk sewer, known as Glattstollen, located in Zurich, Switzerland. This sewer consists of two parallel pipes, which are alternately operated during dry weather conditions (switch every 24 hours) and simultaneously operated during wet weather conditions. There are no lateral confluents, and exfiltration was determined to be excludable due to the concrete construction. The two sewer pipes are approximately 5.3 km in length and have a diameter of 1.1 m. Under dry weather conditions, the travel time is approximately one hour, and the water level is about 0.5 m. It is estimated that the Glattstollen conveys sewage from about 129,000 inhabitants to the main wastewater treatment plant (WWTP) in Zurich, which serves 471,000 inhabitants. This associated subcatchment comprises 149,356 m of sewer pipes, with a wetted area of 50,030 m^2^ under dry weather conditions. All selected respiratory viruses are routinely monitored at the Zurich WWTP (ARA Werdhölzli), along with other sites, as part of the Swiss Wastewater Monitoring Program^4,27^.

### 2.3. Wastewater and biofilm sample collection

Wastewater samples were collected during two sampling campaigns, one in November 2024 and another in January 2025. Wastewater samples were collected at two locations: 0 km (L_0_) and 5 km (L_5_), corresponding to the start and end points of the main trunk sewer. Sampling at L_5_ took place approximately one hour after L_0_ to account for travel time. Twenty-four time-proportional one-hour composite samples were obtained using refrigerated autosamplers that collected a sample every two minutes. Wastewater samples were refrigerated until further analysis.

The biofilm collection campaign was performed in January 2024, during dry weather conditions, in the pipe that was not in operation (approximately two hours after the switch to the other pipe). Biofilm samples were collected in biological triplicates at three distinct sewer locations (i.e., L_0_, L_2.5_, and L_5_) approximately 30 cm above the pipe invert, a position typically submerged over 24 hours under normal flow conditions. An area of 10×10 cm biofilm was collected using a clean spatula and placed in plastic bottles (500 ml) on ice until further analysis. Nine samples were collected in total.

### 2.4. Sample processing and nucleic acid extraction

To assess diurnal viral RNA variations in the bulk liquid, nucleic acids were extracted from individual one-hour composite samples. To evaluate holistic effects of in-sewer transport on viral loads between L_0_ and L_5_, 24-hour composite samples were generated from the one-hour composite samples in a volume-proportional manner for both L_0_ and L_5_. For both assessments, nucleic acids were extracted from a starting volume of 45 mL subsample. One-hour samples were extracted from one subsample, whereas 24-hour composite samples were extracted from three subsamples.

For biofilms, wet biofilm mass was estimated from the bottles’ weights before and after the addition of biofilms. As the extraction kit is optimized for bulk liquid, biofilm samples were re-suspended in phosphate-buffered saline (PBS) to achieve a target biofilm concentration of ∼0.22 g/mL and mixed for

10 min in an orbital shaking incubator (220 rpm). Sample processing for RNA extraction was performed on biofilm samples with a concentration of ∼0.022 g/mL in a total volume of 45 mL (1:10 dilution of original biofilm suspension in PBS).

All nucleic acids were extracted using the Promega Wizard Enviro Total Nucleic Acid Kit (cat. no. A2991) according to manufacturer’s instructions, with some modifications^4^. The Zymo One-Step PCR Inhibitor Removal Kit (cat. no. D6030V) was used to purify extract and reduce PCR inhibition.

### 2.5. RNA recovery efficiency

As an internal control for RNA recovery, approximately 4×10^5^ genome copies (gc) of Murine Hepatitis Virus (MHV) were spiked into all sample types. The samples were incubated for 20 min with orbital shaking at 220 rpm. Following incubation, RNA was extracted and quantified as part of the experiment. To minimize the risk of sample swapping, only a subset of the samples was spiked (i.e., every other sample). RNA extraction efficiency was evaluated by calculating the MHV recovery, expressed as the ratio of measured to expected concentration, presented as a percentage (%).

### 2.6. Digital PCR assay

The RNA quantification of all targets addressed in this study was simultaneously assessed using a previously described six-plex digital PCR (dPCR) assay^4^. This assay targets six gene loci: the SARS-CoV-2 nucleoprotein gene loci N1 and N2, the matrix protein (M) gene of MHV, the M genes of IAV and IBV, and the N gene of RSV. The assay was implemented on the naica® system for the 6-color Crystal Digital PCR (Stilla Technologies, France). Nucleic acid extracts were analyzed in parallel with positive and no-template controls (NTCs). All reactions were performed as previously described. SARS-CoV-2 RNA concentrations were quantified exclusively using N1 target measurements.

To assess potential PCR inhibition, a second round of measurements of the extracted nucleic acids was conducted using synthetic SARS-CoV-2 spiked into the mastermix at approximately 750 gc per reaction. Reactions were also run with only water as template to determine the amount spiked in and the obtained concentration was used to compute the inhibition (expressed in percentage, i.e., ratio of the second round to the first quantification plus amount spiked in). PCR inhibition was considered acceptable when values were below a threshold of 40% as defined according to our internal standard operating procedures^4^. PCR inhibition was assessed in all biofilm extracts and in a selected subset of bulk liquid extracts.

### 2.7. Detection threshold and limit of quantification

Samples were considered positive if their signal reached or exceeded the droplet detection threshold (DT), defined as three positive partitions, which corresponds to approximately 5 genome copies per reaction (gc/rxn). Measurements were deemed quantifiable when they were above the minimum theoretical limit of quantification (LoQ) of 23.25 gc/rxn. This is defined as the concentration at which an ideal dPCR experiment with our assay parameters would yield viral concentration estimates with a coefficient of variation not exceeding 30%.

### 2.8. SARS-CoV-2 sequencing

Nucleic acid extracts from biofilm samples were sequenced to further analyze the quantified amounts of SARS-CoV-2. Sequencing was conducted following Jahn et al.^28^ and processed using V-pipe 3.0^29^. In summary, tiling amplicon PCR amplification was performed with the ARTIC v4.1 protocol^30^. Fragments were sequenced using Illumina Novaseq 6000 250 bp paired-end sequencing. Reads filtering and trimming was performed with PRINSEQ^31^. Alignment to the reference genome (GenBank acc. no. NC_045512.2) was performed using BWA-MEM^32^. Mutation calls were performed using LoFreq^33^. Data were analyzed using Python v3.9. To provide comparative context for the SARS-CoV-2 sequences obtained from biofilm samples, we analyzed mutation profiles in sequencing data from the Zurich WWTP, routinely generated within the framework of the Swiss Wastewater Monitoring Program^28^.

### 2.9. Calculations and statistical analyses

Data analysis was conducted using R (v4.1.1) and RStudio (v2024.9.1.394). Viral RNA concentrations in nucleic acid extracts were calculated using Crystal Miner software (v4.0) from Stilla Technologies and are reported as genome copies per microliter of reaction (gc/µL of rxn). Depending on the sample type, concentrations were further expressed as genome copies per liter of wastewater (gc/L_ww_) for bulk liquid samples, or genome copies per gram (gc/g) for biofilm samples. Viral RNA loads are calculated by multiplying concentrations with wastewater flow rates.

The R package “dPCRfit” was used to estimate the percentage reduction in viral RNA concentrations between sewer locations^34^. For each viral target and sampling experiment, a Bayesian generalized linear model (GLM) was fitted with a logarithmic link to the three biological replicates measured at L_0_ and L_5_, respectively, using the sewer location as an independent variable. The GLM used a dPCR-specific likelihood tailored to the parameters of our assay^35^. We report the median and 95% credible intervals (CrIs) of the estimated percentage change in expected viral concentration between L_0_ and L_5_ (additional details are provided in **Text S1**).

## 3. Results

### 3.1. Diurnal patterns in viral RNA levels

We conducted the experiments during two experimental campaigns, on 7-8 November 2024 and 20-21 January 2025. In November 2024, SARS-CoV-2 was the predominant pathogen in circulation, while other targeted pathogens were circulating at much lower levels^27^. The autosampler was initiated at approximately 09:00 and run for 24 hours. One-hour composite samples were then analyzed individually, and concentrations were measured for each sample (**Figure S1**). SARS-CoV-2 was quantifiable in all 24 one-hour composite samples, whereas RSV was quantifiable in only two samples. Both IAV and IBV were either undetectable or below the limit of quantification (LoQ). In January 2025, the autosampler was started at approximately 11:00, and all targeted viruses were circulating during this period, resulting in more samples with elevated concentrations (**Figure S2**). Indeed, all pathogens were detectable in every sample, with only a few cases where some were below the LoQ.

To enable comparison between the two sampling experiments despite differences in concentrations and starting times, sampling times were adjusted to a common timeline from 00:00 to 23:00 (**Figure 1**). Viral RNA levels in wastewater exhibited a reproducible, systematic diurnal pattern, consistently peaking between 7:00 and 11:00 across all tested pathogens and experiments. During these four hours (1/6^th^ of the day), the viral load accounted for a substantial proportion of the total daily viral load, with variations observed depending on the experiment and target. Specifically, for SARS-CoV-2, the viral load during these four hours accounted for 50% (November 2024 experiment) and 45% (January 2025 experiment) of the total daily load, for IAV 47% and 32%, for IBV 50% and 22%, and for RSV 41% and 34%.

**Figure 1.**
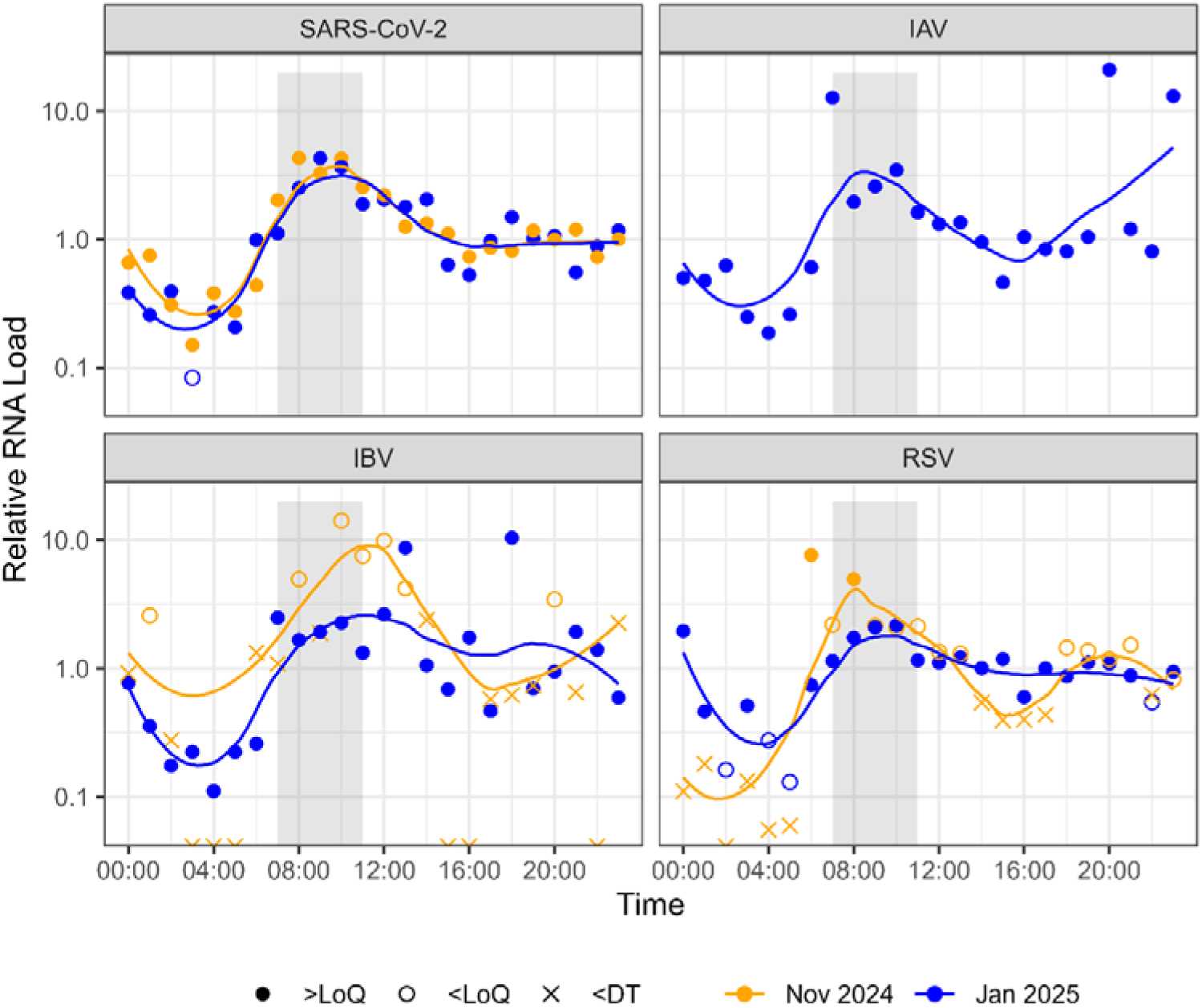
Diurnal variations in viral RNA loads. The vertical axis represents the relative RNA load and it is shown in log_10_-scale. The horizontal axis indicates the time of day. Data points represent individual measurements, with orange points for the November 2024 experiment and blue points for the January 2025 experiment. Symbols shape describes if measurements are below the detection threshold (DT; cross), below LoQ (empty circle), or above LoQ (filled circle). Solid lines display smoothed trends using the LOESS method. Each panel corresponds to a different respiratory virus. To facilitate comparison between the two experiments and to align diurnal patterns, sampling times were adjusted to a common 00:00-23:00 timeline. IAV: Influenza A Virus. IBV: Influenza B Virus. RSV: Respiratory Syncytial Virus.

### 3.2. Influence of in-sewer transport on viral RNA concentrations

We investigated the changes in viral RNA concentrations in a trunk sewer without exfiltration and no lateral confluents, over a length of 5 km (from L_0_ to L_5_), with an approximate travel time of one hour. Our experiments aimed to assess how viral RNA concentrations may be impacted as wastewater travels through the sewer. The current experiment included two sampling campaigns, conducted on 26-27 November 2024 and 21-22 January 2025. Viral concentrations were consistently higher during the January campaign. In November 2024, by contrast, all viruses exhibited concentrations below LoQ, and in some instances, below the DT in the 24-hour composite samples, except for SARS-CoV-2 (**Figure 2**).

**Figure 2.**
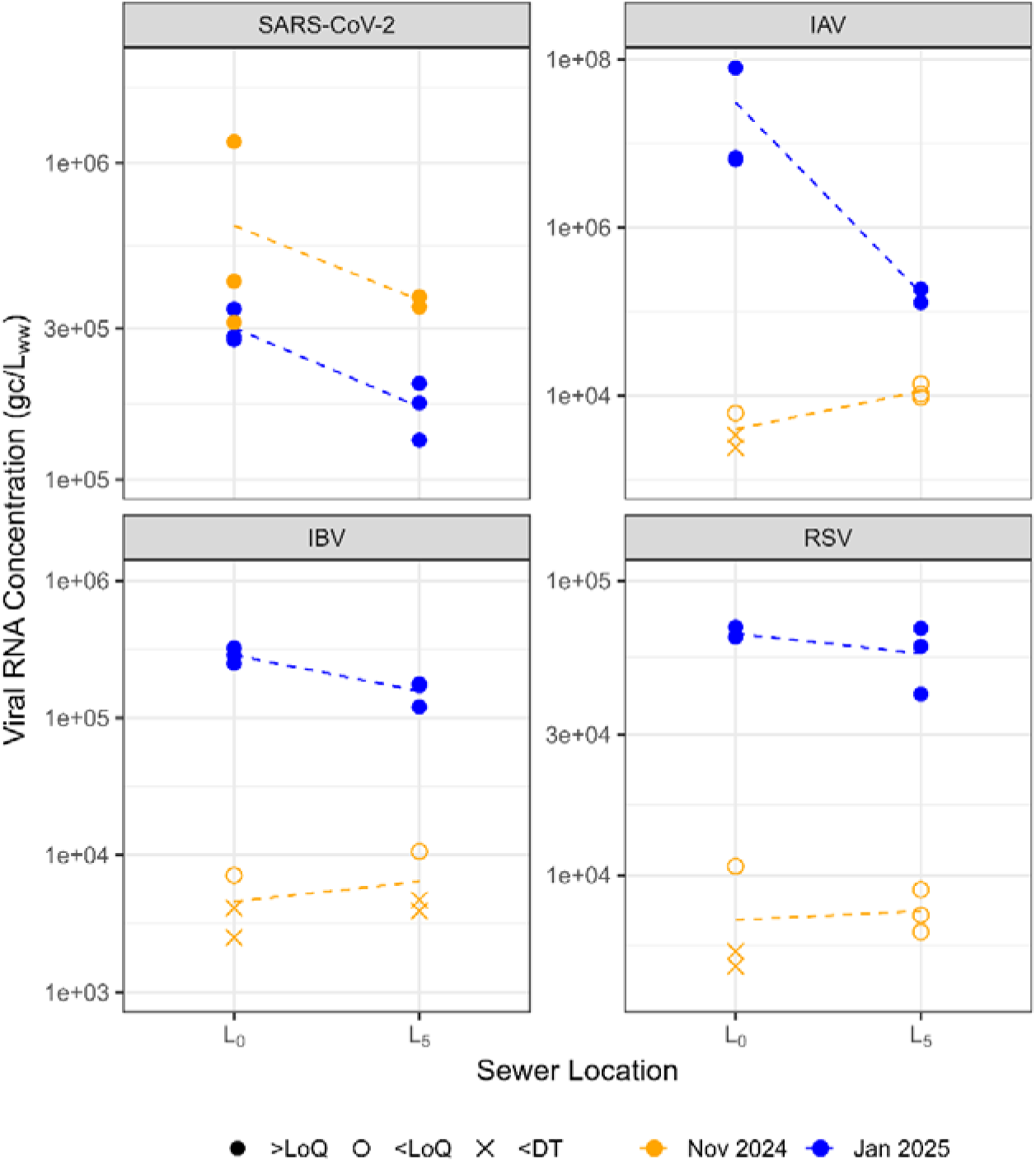
Viral RNA concentration changes in a sewer pipe of 5 km length and one hour of travel time. The vertical axes represent the viral RNA concentration, expressed in genome copies (gc) per liter of wastewater (L_ww_), while the horizontal axes indicate the locations in the sewer pipe at 0 km (L_0_) and at 5 km (L_5_). Individual data points represent measurements from biological extraction replicates. Orange symbols correspond to the November 2024 experiment, while blue symbols represent the January 2025 experiment. Filled circles denote values above the theoretical minimum limit of quantification (LoQ), empty circles indicate values below LoQ but above the detection threshold (DT), and crosses represent measurements below the DT. Each panel corresponds to a different respiratory virus (IAV: Influenza A Virus, IBV: Influenza B Virus, RSV: Respiratory Syncytial Virus).

To highlight the differences between L_0_ and L_5_, we chose to focus on data from the January 2025 experiment, as RNA concentrations in the samples were predominantly above the LoQ during this period. We observed a change in pathogen levels in the bulk liquid between the two sewer locations, with the following estimated median percent changes: -43% (95% credible interval [CrI]: -58%, -22%) for SARS-CoV-2, -72% (CrI: -96%, 64%) for IAV, -44% (CrI: -60%, -22%) for IBV, and -15% (CrI: -36%, 20%) for RSV (**Figure S3**). For SARS-CoV-2, which exhibited quantifiable RNA levels in November 2024 as well, the estimated median percent change was comparable (-37%, 95% CrI: - 68%, 42%); however, this difference did not reach statistical significance due to the high variability observed at L_0_ (coefficient of variation: 73%).

In both sampling campaigns, we measured total suspended solids (TSS) in 24-hour composite samples. Between locations L_0_ and L_5_, TSS concentrations decreased from 328 mg/L to 130 mg/L (60%) in November and from 392 mg/L to 158 mg/L (60%) in January. PCR inhibition was evaluated and showed no significant differences between L_0_ and L_5_ in either sampling campaign (t-test; *p* = 0.29 and 0.25, respectively).

In this study, we also assessed RNA recovery efficiency from bulk liquid samples using Murine Hepatitis Virus (MHV) as a surrogate control. Out of 60 total nucleic acid extractions performed, a subset of 32 wastewater samples was spiked with a known concentration of MHV prior to concentration and extraction. Our analysis revealed a mean RNA recovery of 33% ± 4%.

### 3.3. Influence of sewer biofilms on viral RNA concentrations

In January 2024, sewer biofilm samples were collected in biological triplicates from three distinct locations along the sewer system: L_0_, L_2.5_, and L_5_. The wet biofilm mass was consistent across triplicate samples at each location, as indicated by low to moderate coefficients of variation: 16% at L_0_ (mean ± SD: 17.3 ± 2.7 g), 3% at L_2.5_ (19.4 ± 0.6 g), and 13% at L_5_ (9.2 ± 1.2 g). However, the wet mass varied significantly with distance (*p* = 0.0008, ANOVA). The mass at L_5_ was notably lower than at both L_0_ (*p* = 0.003, Tukey HSD) and L_2.5_ (*p* = 0.0009), whereas no difference was observed between L_0_ and L_2.5_ (*p* = 0.34).

SARS-CoV-2 and IAV were detected in all nine biofilm samples, while RSV was present in seven out of nine samples and IBV in two samples (**Figure 3**). For RSV and IBV, concentrations were generally low, below the LoQ, reflecting the low prevalence of these viruses during the sampling period in January 2024.

**Figure 3.**
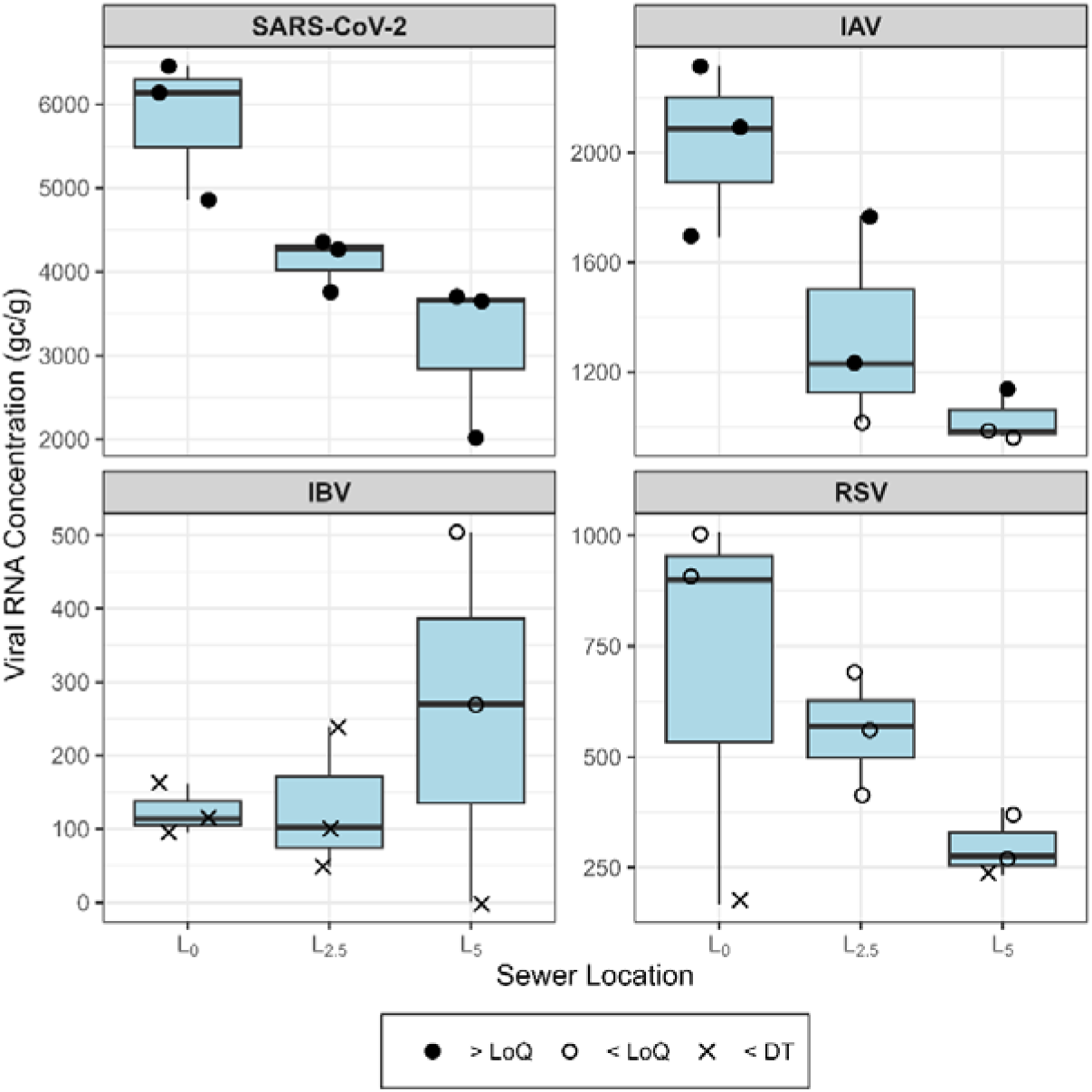
Viral RNA concentration of common respiratory viruses in sewer biofilms. The vertical axes indicate viral RNA concentrations expressed in genome copies (gc) per gram (g) of wet biofilm. The horizontal axes describe the sewer location. Data is presented with boxplots, individual measurements are shown with symbols: filled circles represent values above the limit of quantification (LoQ), empty circles indicate values below LoQ, and crosses indicate values below the detection threshold (DT). Each panel corresponds to a different respiratory virus (IAV: Influenza A Virus, IBV: Influenza B Virus, RSV: Respiratory Syncytial Virus).

SARS-CoV-2 RNA was detected in all samples, with median concentrations of 6,132 genome copies (gc)/g of wet biofilm at L_0_, 4,272 gc/g at L_2.5_, and 3,660 gc/g at L_5_. RNA concentrations were 46% lower at L_5_ compared to L_0_ (*p* = 0.01), which was in line with the reduction observed for the bulk liquid. All samples also tested positive for IAV RNA. Median concentrations decreased from 2,088 gc/g at L_0_ to 1,230 gc/g at L_2.5_ and 984 gc/g at L_5_. This corresponded to a 49% reduction at L_5_ relative to L_0_ (*p* = 0.01). Importantly, no difference in MHV recovery was present between samples collected at different sewer locations (*p* = 0.56). However, overall recovery efficiency was relatively low, averaging 8% ± 1%. PCR inhibition also did not differ among collecting locations (*p* = 0.06).

The Swiss Wastewater Monitoring Program routinely quantifies viral RNA loads at the Zurich WWTP. At the time of biofilm sampling (17 January 2024), the total SARS-CoV-2 load in the catchment, estimated using measurements from 16 January 2024, was 6.46×10^13^ gc. Since the population served by the main trunk sewer of interest represents approximately 27% of the total catchment population, we estimated that the corresponding viral load in the analyzed subcatchment was approximately 1.75×10^13^ gc. Using data on subcatchment characteristics, we estimated the SARS-CoV-2 RNA retained in the total sewer biofilm. This was calculated by multiplying the average RNA concentration in the biofilm (gc/g) by the biofilm’s mass per unit area (g/m^2^) and the wetted surface area of the sewers, as provided by the operators of the sewer system based on a hydrodynamic model. Assuming no in-sewer RNA loss, our results indicate that the biofilm-associated RNA accounted for approximately 2% of the total viral load in the subcatchment, which includes both viral RNA in the bulk wastewater and that associated with the biofilm matrix.

### 3.4. Whole genome sequencing of SARS-CoV-2 from biofilm samples

Since SARS-CoV-2 RNA was quantifiable in the biofilm samples, we also performed whole genome sequencing to explore the genetic composition in more detail. We first analyzed the genome coverage and per-position read depth to assess sequencing quality. Most samples failed to achieve sufficient coverage for reliable downstream analysis, except for two samples (third replicate at 0 km length, L_0__3 and first replicate at 5 km length, L_5__1) (**Figure S4**). Next, we performed mutation calling on both biofilm and influent wastewater samples from the Zurich WWTP to contextualize the mutations identified in the biofilm (**Figure S5**). The mutation profiles observed in samples L_0__3 and L_5__1 were most strongly correlated with those from WWTP samples temporally closest to the biofilm sampling date (17 January 2024; **Figure S6**). Indeed, the correlation between the WWTP samples and the biofilm samples rises starting September 2023, and plateaus starting mid-December 2023, and we observed the highest correlation coefficients, with r = 0.96 for L_0__3 on 13 January 2024 and r = 0.90 for L_5__1 on 5 January 2024, respectively.

## 4. Discussion

This study provides insights into viral RNA intra-day dynamics and potential losses during in-sewer transport through a series of field experiments conducted in a real-world sewer. The experiments in our subcatchment revealed that the concentrations and loads of four selected viral pathogens in the bulk liquid exhibit a clear diurnal pattern, with peak levels observed in the morning. This trend likely reflects daily routines such as morning toilet use, suggesting that viruses, similar to other substances, are excreted primarily during the morning hours^36^. Observations of diurnal patterns also imply that wastewater travels quickly from households to downstream locations, as longer residence times and wider travel time distributions would result in increased dispersion, which would level out temporal variations in loads. This diurnal variability highlights a practical tradeoff. If the goal is to ensure consistency in tracking viral signals and to generate reliable longitudinal time series for monitoring, 24-hour composite sampling seems the best solution, since it covers the entire day and it is resistant to fluctuations in diurnal patterns (as might be expected between weekdays and weekends due to different routine habits, such as waking times). However, if the objective is early detection of emerging diseases, collecting composite samples during specified morning hours only, when viral concentrations are highest, may improve sensitivity and enhance monitoring efforts. But this approach may also increase bias toward communities with shorter residence times to the sampling location compared with 24-hour composite sampling.

To investigate the impact of in-sewer transport on viral RNA estimates, we analyzed changes in viral concentrations between two sampling locations along a trunk sewer, spaced approximately 5 km apart with an estimated wastewater travel time of one hour (L_0_ to L_5_). We observed variable reductions in viral RNA concentrations between the two locations, with estimated percentage changes ranging from 15% to 72% across different viral targets. Notably, for SARS-CoV-2, which was consistently detectable with relatively high confidence during both sampling campaigns, the percentage reduction and the rate of signal loss were consistent across experiments. This consistency was observed regardless of the initial viral concentration at the upstream site (6.3×10^5^ gc/L_ww_ in November 2024 and 3.0×10^5^ gc/L_ww_ in January 2025), suggesting that the pattern of signal attenuation during in-sewer transport remains stable over time. This interpretation is further supported by the observation that diurnal fluctuation patterns were also similar across two separate sampling periods conducted in different months. Nonetheless, the experiments were conducted only twice, and the observed patterns may differ in other locations or under different flow regimes.

The observed reduction in viral RNA concentrations suggests substantial signal loss during in-sewer transport. This loss is likely driven by multiple factors. While RNA decay occurs in the wastewater matrix, the main drivers were reported to be pH and temperature^37^. Given that these factors are expected to be rather stable over the short time frame (i.e., one hour), and that even under conditions promoting RNA degradation substantial decay typically requires longer durations^38^, it is rather unlikely that RNA decay alone accounts for the observed extent of reduction.

Numerous studies have demonstrated that viral RNA is highly associated with solids^2,38–40^, either because it is mainly excreted in feces already bound to particulate matter or because it adsorbs to suspended solids during in-sewer transport. Therefore, the loss of solids through sedimentation or settling can lead to a reduction of viral RNA in the bulk liquid. To support this hypothesis, we examined total suspended solids between L_0_ and L_5_ and observed a measurable reduction, suggesting that sedimentation may contribute to the observed signal loss, despite the sustained flow rate of approximately 0.98 m^3^/s in November and 0.65 m^3^/s in January. The association between viral RNA and solids explains why some wastewater surveillance programs focus on extracting viral RNA from the solids rather than the liquid phase or the bulk liquid^41^. However, substantial sedimentation implies that a variable fraction of solids may be lost before sampling^42^.

Viral RNA adsorption can also take place onto biofilms that adhere to the sewer walls. Our findings suggest that sewer biofilms act as a minor reservoir of viral RNA. Indeed, the proportion of SARS-CoV-2 RNA estimated to be sorbed to biofilms across the entire catchment, approximately 2% of the total RNA, differs substantially from the value reported by Li et al., which showed that intact biofilms grown in a controlled reactor system retained up to 78% of SARS-CoV-2 RNA^17^. This discrepancy may be attributed to several factors. Compared with sewers, reactors operate at lower flow velocities and longer retention times and provide substantially greater effective surface area, as well as more stable physicochemical conditions. Together, these characteristics might enhance virus-surface interactions and promote adsorption. Separately, our nucleic acid extraction protocol was not specifically optimized for solids, which could have limited RNA recovery from the biofilms. Although recovery was relatively low when assessed using a surrogate as control, it was similar at the collected biofilm locations, and we still observed a substantial decrease in viral concentrations in the biofilm along the sewer stretch, reinforcing the observation that the viral signal is progressively lost rather than reflecting poorer recovery at one sampling location than another.

As sewer biofilms act as reservoirs, this can lead to adsorption-desorption kinetics that attenuate viral signals in wastewater. SARS-CoV-2 variants detected in bulk liquid may therefore originate from biofilms sloughing off, rather than reflecting active circulation among infected individuals. In our study, the sequence variation in biofilms closely matched that in temporally coincident bulk water, indicating that the SARS-CoV-2 population in biofilms was not distinct from that in the surrounding bulk water, and that SARS-CoV-2 RNA adsorbed to biofilms is only short term preserved.

According to our findings, the observed extent of RNA reduction over a 5-km stretch suggests that catchments with longer residence times may exhibit estimated lower viral loads when measured as raw influent to the WWTP. This has important implications, as it could lead to underestimation of viral loads and pose challenges for models aiming to predict disease prevalence from wastewater data. In particular, it underscores the need of catchment-specific relationships between measured viral loads and actual disease incidence. Accounting for travel-time-dependent RNA loss could help resolve spatial contributions to wastewater measurements and reveal finer patterns in clinical incidence. With additional sampling points and a well-characterized sewer network, observed viral concentrations could be modeled as upstream inputs weighted by transport, enabling estimation of subcatchment-level viral loads. This approach currently involves substantial uncertainty due to limited data but could become more robust with additional sampling experiments and improved network characterization.

Although not the primary focus of our study, we also highlight another substantial potential source of signal loss, which is RNA recovery efficiency during extraction. Using a viral surrogate, we estimated an average loss of 67%, however, this surrogate is not necessarily representative of all circulating viruses^43^. Inhibition of samples could also play a role in loss of signal, however in our samples, we did not observe significant impact of inhibition on estimated concentrations from upstream to downstream sites.

## 5. Conclusions

Overall, this study provides a foundation for future research on RNA dynamics in wastewater and underscores the complexity of in-sewer transport processes. Moreover, our observations emphasize that advancing toward mechanistic models of disease incidence, rather than relying solely on empirical correlations, will not only require more precise, quantitative insights into how viral signals fluctuate and attenuate within sewer networks, but also depend on further real-world field experiments across diverse sewer systems worldwide.

## Data Availability

All data produced in the present study are available upon reasonable request to the authors.

## Author Contributions

MP: Conceptualization; Data curation; Formal analysis; Investigation; Methodology; Visualization; Writing - original draft. CG: Methodology; Writing - review & editing. SB: Methodology; Writing - review & editing. DD: Formal analysis; Investigation; Visualization; Writing - review & editing. AL: Formal analysis; Investigation; Visualization; Writing - review & editing. TRJ: Funding acquisition; Supervision; Writing - review & editing. CO: Conceptualization; Funding acquisition; Methodology; Supervision; Writing - review & editing.

## Acknowledgements

We gratefully acknowledge the support of the Entsorgung + Recycling Zürich (ERZ) team in this study. In particular, we thank Kai Hübscher, Roger Küenzi, Domenico Sansone, Rey Eyer, and Niculin Cathomen for their valuable assistance. We also thank Sylvia Richter and Fabian Kuhn for their technical support in quantifying total suspended solids in the wastewater samples. We thank past and present members of the Wastewater Monitoring Laboratory team at Eawag for their assistance with sample processing, and collaborators of the WISE (Wastewater-based Infectious disease Surveillance and Epidemiology) project for their support and contributions. We also thank NEXUS Personalized Health Technologies for their support with the bioinformatic analysis.

## References

1 Hopkins, L. et al. Citywide wastewater SARS-CoV-2 levels strongly correlated with multiple disease surveillance indicators and outcomes over three COVID-19 waves. Science of The Total Environment 855, 158967 (2023). 10.1016/j.scitotenv.2022.158967

2 Graham, K. E. et al. SARS-CoV-2 RNA in Wastewater Settled Solids Is Associated with COVID-19 Cases in a Large Urban Sewershed. Environmental Science & Technology 55, 488–498 (2021). 10.1021/acs.est.0c06191

3 Bagutti, C. et al. Wastewater monitoring of SARS-CoV-2 shows high correlation with COVID-19 case numbers and allowed early detection of the first confirmed B.1.1.529 infection in Switzerland: results of an observational surveillance study. Swiss Medical Weekly 152, w30202 (2022). 10.4414/SMW.2022.w30202

4 Pitton, M. et al. A six-plex digital PCR assay for monitoring respiratory viruses in wastewater. Nature Water 3, 1174–1186 (2025). 10.1038/s44221-025-00503-x

5 Li, X., Zhang, S., Shi, J., Luby, S. P. & Jiang, G. Uncertainties in estimating SARS-CoV-2 prevalence by wastewater-based epidemiology. Chemical Engineering Journal 415, 129039 (2021). 10.1016/j.cej.2021.129039

6 Wade, M. J. et al. Understanding and managing uncertainty and variability for wastewater monitoring beyond the pandemic: Lessons learned from the United Kingdom national COVID-19 surveillance programmes. Journal of Hazardous Materials 424, 127456 (2022). 10.1016/j.jhazmat.2021.127456

7 Li, T.-Z. et al. Duration of SARS-CoV-2 RNA shedding and factors associated with prolonged viral shedding in patients with COVID-19. Journal of Medical Virology 93, 506–512 (2021). 10.1002/jmv.26280

8 Zhou, B., She, J., Wang, Y. & Ma, X. Duration of Viral Shedding of Discharged Patients With Severe COVID-19. Clinical Infectious Diseases 71, 2240–2242 (2020). 10.1093/cid/ciaa451

9 Cevik, M. et al. SARS-CoV-2, SARS-CoV, and MERS-CoV viral load dynamics, duration of viral shedding, and infectiousness: a systematic review and meta-analysis. The Lancet Microbe 2, e13–e22 (2021). 10.1016/S2666-5247(20)30172-5

10 Crank, K., Chen, W., Bivins, A., Lowry, S. & Bibby, K. Contribution of SARS-CoV-2 RNA shedding routes to RNA loads in wastewater. Science of The Total Environment 806, 150376 (2022). 10.1016/j.scitotenv.2021.150376

11 Jones, D. L. et al. Shedding of SARS-CoV-2 in feces and urine and its potential role in person-to-person transmission and the environment-based spread of COVID-19. Science of The Total Environment 749, 141364 (2020). 10.1016/j.scitotenv.2020.141364

12 Amoah, I. D. et al. Effect of selected wastewater characteristics on estimation of SARS-CoV-2 viral load in wastewater. Environmental Research 203, 111877 (2022). 10.1016/j.envres.2021.111877

13 Burnet, J.-B., Cauchie, H.-M., Walczak, C., Goeders, N. & Ogorzaly, L. Persistence of endogenous RNA biomarkers of SARS-CoV-2 and PMMoV in raw wastewater: Impact of temperature and implications for wastewater-based epidemiology. Science of The Total Environment 857, 159401 (2023). 10.1016/j.scitotenv.2022.159401

14 Breadner, P. R. et al. A comparative analysis of the partitioning behaviour of SARS-CoV-2 RNA in liquid and solid fractions of wastewater. Science of The Total Environment 895, 165095 (2023). 10.1016/j.scitotenv.2023.165095

15 Ye, Y., Ellenberg, R. M., Graham, K. E. & Wigginton, K. R. Survivability, Partitioning, and Recovery of Enveloped Viruses in Untreated Municipal Wastewater. Environmental Science & Technology 50, 5077–5085 (2016). 10.1021/acs.est.6b00876

16 Roldan-Hernandez, L. & Boehm, A. B. Adsorption of Respiratory Syncytial Virus, Rhinovirus, SARS-CoV-2, and F+ Bacteriophage MS2 RNA onto Wastewater Solids from Raw Wastewater. Environmental Science & Technology 57, 13346–13355 (2023). 10.1021/acs.est.3c03376

17 Li, J. et al. Impact of sewer biofilms on fate of SARS-CoV-2 RNA and wastewater surveillance. Nature Water 1, 272–280 (2023). 10.1038/s44221-023-00033-4

18 Ort, C., Lawrence, M. G., Reungoat, J. & Mueller, J. F. Sampling for PPCPs in Wastewater Systems: Comparison of Different Sampling Modes and Optimization Strategies. Environmental Science & Technology 44, 6289–6296 (2010). 10.1021/es100778d

19 Qiu, J. Y. et al. Impact of Sample Storage Time and Temperature on the Stability of Respiratory Viruses and Enteric Viruses in Wastewater. Microorganisms 12, 2459 (2024). 10.3390/microorganisms12122459

20 Tavazzi, S. et al. Short-term stability of wastewater samples for storage and shipment in the context of the EU Sewage Sentinel System for SARS-CoV-2. Journal of Environmental Chemical Engineering 11, 109623 (2023). 10.1016/j.jece.2023.109623

21 Zheng, X. et al. Comparison of virus concentration methods and RNA extraction methods for SARS-CoV-2 wastewater surveillance. Science of The Total Environment 824, 153687 (2022). 10.1016/j.scitotenv.2022.153687

22 Jiang, M. et al. Evaluation of the Impact of Concentration and Extraction Methods on the Targeted Sequencing of Human Viruses from Wastewater. Environmental Science & Technology 58, 8239–8250 (2024). 10.1021/acs.est.4c00580

23 O’Brien, M. et al. A comparison of four commercially available RNA extraction kits for wastewater surveillance of SARS-CoV-2 in a college population. Science of The Total Environment 801, 149595 (2021). 10.1016/j.scitotenv.2021.149595

24 Ahmed, W. et al. Comparison of RT-qPCR and RT-dPCR Platforms for the Trace Detection of SARS-CoV-2 RNA in Wastewater. ACS ES&T Water 2, 1871–1880 (2022). 10.1021/acsestwater.1c00387

25 Michael-Kordatou, I., Karaolia, P. & Fatta-Kassinos, D. Sewage analysis as a tool for the COVID-19 pandemic response and management: the urgent need for optimised protocols for SARS-CoV-2 detection and quantification. Journal of Environmental Chemical Engineering 8, 104306 (2020). 10.1016/j.jece.2020.104306

26 Toze, S. PCR and the detection of microbial pathogens in water and wastewater. Water Research 33, 3545–3556 (1999). 10.1016/S0043-1354(99)00071-8

27 Wastewater-based Infectious Disease Surveillance and Epidemiology Dashboard, <https://wise.ethz.ch/> (2025). Accessed: 5 May 2025.

28 Jahn, K. et al. Early detection and surveillance of SARS-CoV-2 genomic variants in wastewater using COJAC. Nature Microbiology 7, 1151–1160 (2022). 10.1038/s41564-022-01185-x

29 Fuhrmann, L. et al. V-pipe 3.0: a sustainable pipeline for within-sample viral genetic diversity estimation. GigaScience 13 (2024). 10.1093/gigascience/giae065

30 DNA Pipelines R&D et al. COVID-19 ARTIC v3 Illumina library construction and sequencing protocol. protocols.io (2020). 10.17504/protocols.io.bgxjjxkn

31 Schmieder, R. & Edwards, R. Quality control and preprocessing of metagenomic datasets. Bioinformatics 27, 863–864 (2011). 10.1093/bioinformatics/btr026

32 Li, H. & Durbin, R. Fast and accurate short read alignment with Burrows–Wheeler transform. Bioinformatics 25, 1754–1760 (2009). 10.1093/bioinformatics/btp324

33 Wilm, A. et al. LoFreq: a sequence-quality aware, ultra-sensitive variant caller for uncovering cell-population heterogeneity from high-throughput sequencing datasets. Nucleic Acids Research 40, 11189–11201 (2012). 10.1093/nar/gks918

34 Lison, A. dPCRfit, <https://github.com/adrian-lison/dPCRfit> (2025). Accessed: 19 May 2025.

35 Lison, A., Julian, T. R. & Stadler, T. Improving inference in environmental surveillance by modeling the statistical features of digital PCR. bioRxiv, 2024.2010.2014.618307 (2025). 10.1101/2024.10.14.618307

36 Coutu, S., Wyrsch, V., Wynn, H. K., Rossi, L. & Barry, D. A. Temporal dynamics of antibiotics in wastewater treatment plant influent. Science of The Total Environment 458-460, 20–26 (2013). 10.1016/j.scitotenv.2013.04.017

37 Korajkic, A., McMinn, B. R., Pemberton, A. C., Kelleher, J. & Ahmed, W. The comparison of decay rates of infectious SARS-CoV-2 and viral RNA in environmental waters and wastewater. Science of The Total Environment 946, 174379 (2024). 10.1016/j.scitotenv.2024.174379

38 Zhang, M., Roldan-Hernandez, L. & Boehm, A. Persistence of human respiratory viral RNA in wastewater-settled solids. Applied and Environmental Microbiology 90, e02272–02223 (2024). 10.1128/aem.02272-23

39 Kitamura, K., Sadamasu, K., Muramatsu, M. & Yoshida, H. Efficient detection of SARS-CoV-2 RNA in the solid fraction of wastewater. Science of The Total Environment 763, 144587 (2021). 10.1016/j.scitotenv.2020.144587

40 D’Aoust, P. M. et al. Quantitative analysis of SARS-CoV-2 RNA from wastewater solids in communities with low COVID-19 incidence and prevalence. Water Research 188, 116560 (2021). 10.1016/j.watres.2020.116560

41 Boehm, A. B. et al. Human viral nucleic acids concentrations in wastewater solids from Central and Coastal California USA. Scientific Data 10, 396 (2023). 10.1038/s41597-023-02297-7

42 Nourbakhsh, S. et al. Mechanistic modelling of in-sewer viral fate and transport of SARS-CoV-2 to enhance wastewater disease surveillance strategies. PREPRINT (Version 1) available at Research Square (2025). 10.21203/rs.3.rs-7774650/v1

43 Kantor, R. S., Nelson, K. L., Greenwald, H. D. & Kennedy, L. C. Challenges in Measuring the Recovery of SARS-CoV-2 from Wastewater. Environmental Science & Technology 55, 3514–3519 (2021). 10.1021/acs.est.0c08210

