## Supplementary Material for "Tracking viral RNA loads during in-sewer transport: insights from real-world field experiments"

**Supplementary Figures**

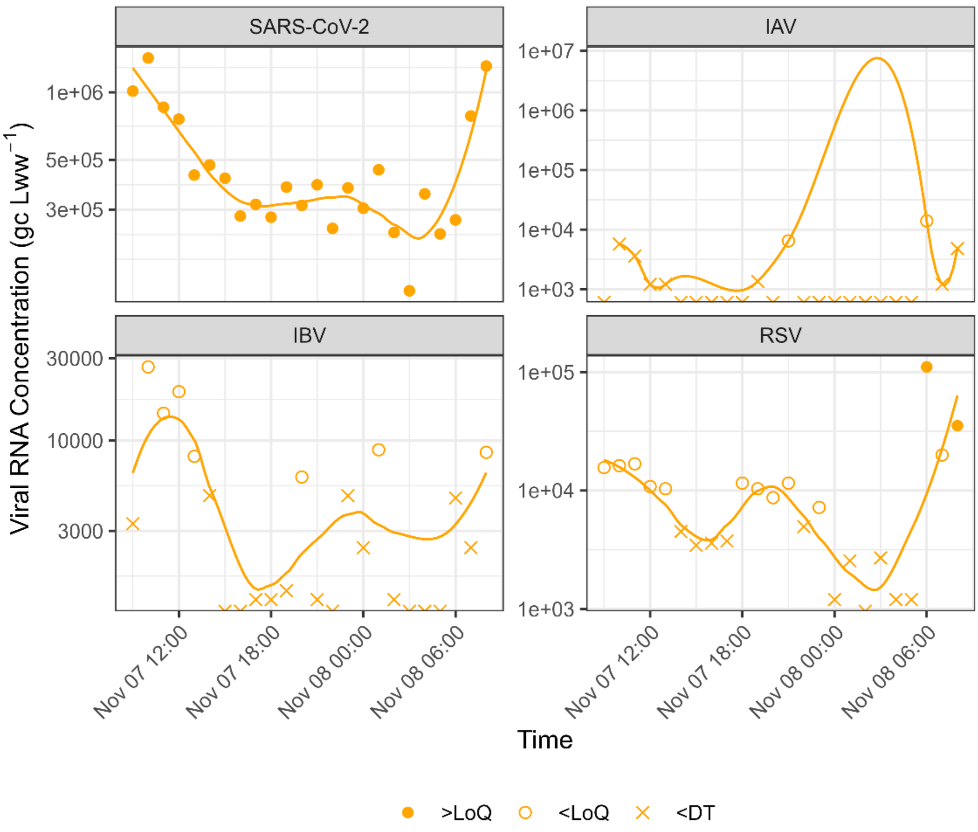


**Figure S1. Viral RNA concentration profile over a 24-hour period in November 2024.** The vertical axis represents the concentration of viral RNA expressed in genome copies (gc) per liter of wastewater ($L_{ww}$). The horizontal axis indicates the date and time of day. Data points represent individual measurements. Symbols shape describes if measurements are above LoQ (filled circle), below LoQ (empty circle), or below DT (cross). Solid lines display smoothed trends using the loess method (span = 0.5). Each panel corresponds to a different respiratory virus. IAV: Influenza A Virus; IBV: Influenza B Virus; RSV: Respiratory Syncytial Virus.

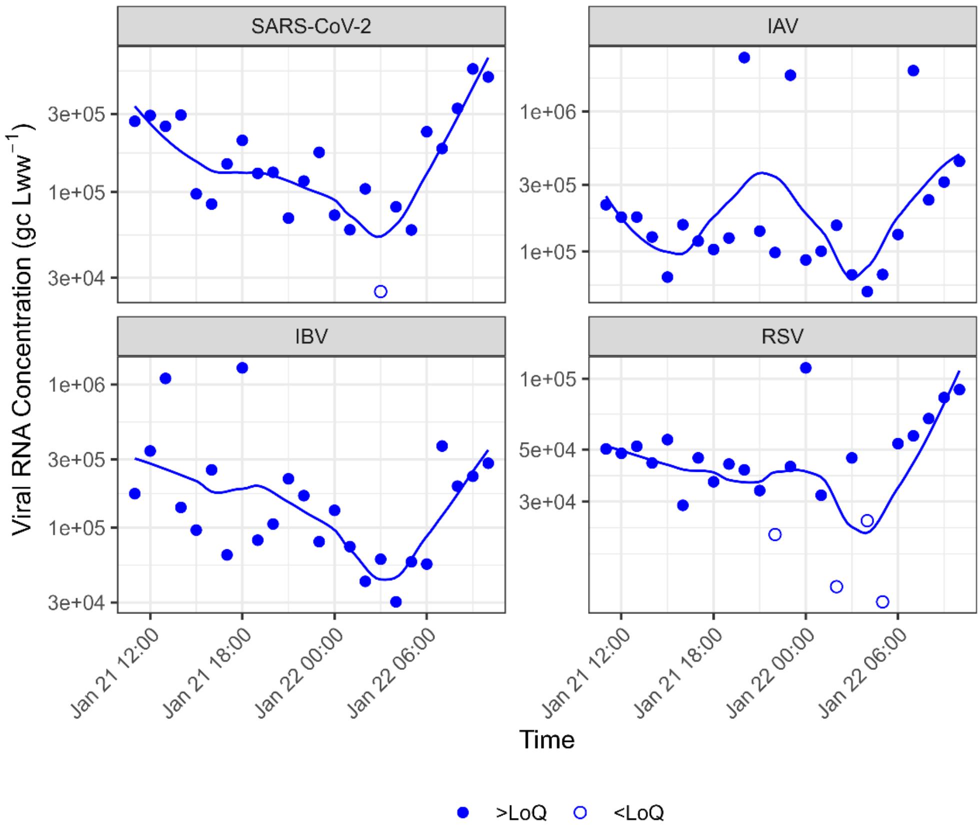


**Figure S2. Viral RNA concentration profile over a 24-hour period in January 2025.** The vertical axis represents the concentration of viral RNA expressed in genome copies (gc) per liter of wastewater ($L_{ww}$). The horizontal axis indicates the date and time of day. Data points represent individual measurements. Symbols shape describes if measurements are above LoQ (filled circle) or below LoQ (empty circle). Solid lines display smoothed trends using the loess method (span = 0.5). Each panel corresponds to a different respiratory virus. IAV: Influenza A Virus; IBV: Influenza B Virus; RSV: Respiratory Syncytial Virus.

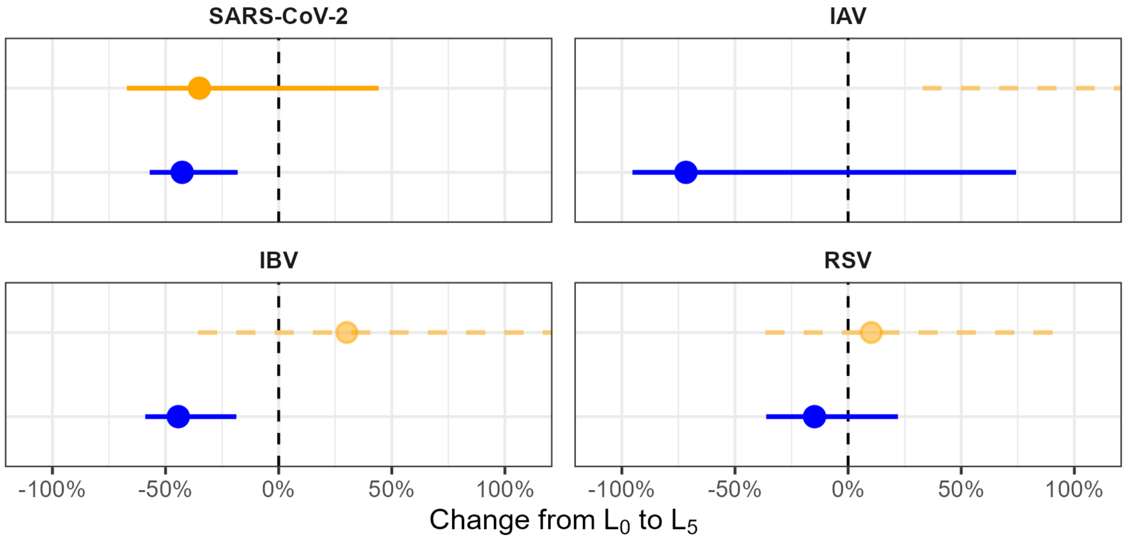


**Figure S3. Percentage change in viral RNA concentrations between locations $L_0$ and $L_5$.** The horizontal axis shows the median estimated change (filled circle) with associated confidence intervals (horizontal bars) for the two sampling experiments (orange: November 2024, blue: January 2025). Values to the left of the dashed vertical line (0%) indicate a decrease in concentration along the sewer, while values to the right indicate an increase. Slightly transparent filled circles with dashed horizontal bars represent data below the limit of quantification.

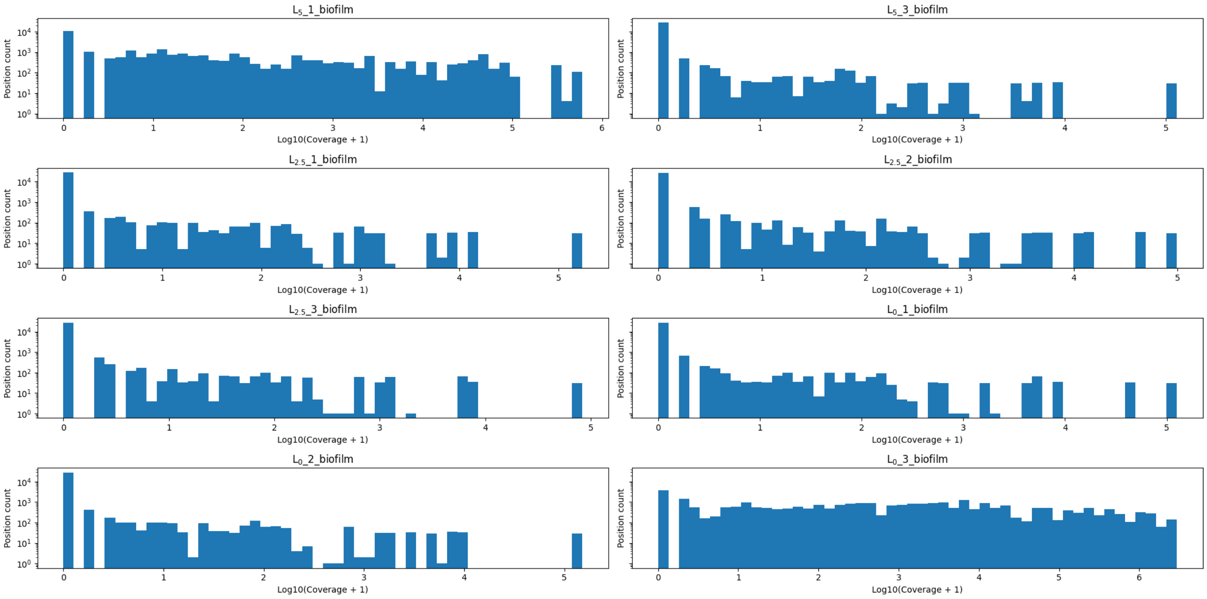


**Figure S4. Distribution of coverage across genomic regions for biofilm samples.** The horizontal axes show the $log_{10}$ read depth (with an added pseudocount), and vertical axes show the count of position with that read depth. Each panel refers to a different biofilm sample (2 replicates at $L_5$, 3 replicates at $L_{2.5}$ and 3 replicates at $L_0$).

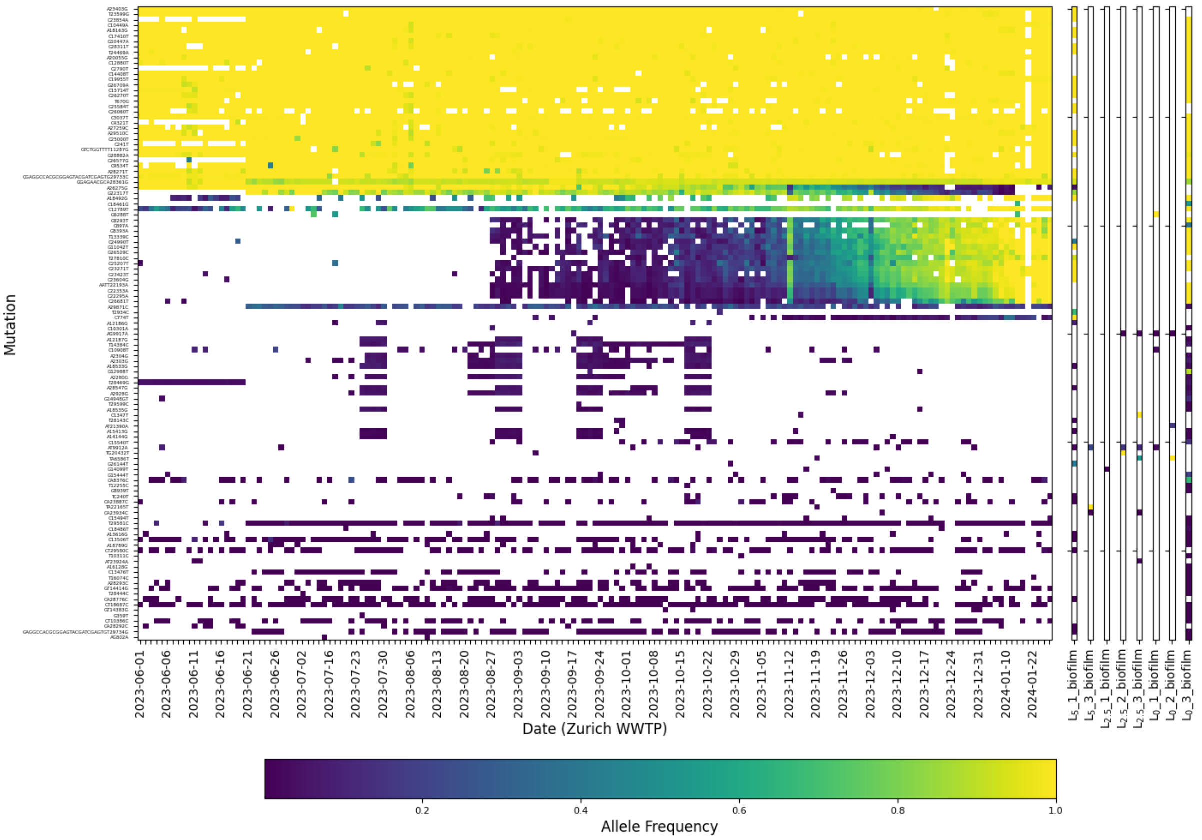


**Figure S5. Heatmap of allele frequency of mutations detected in biofilm samples.** The left panel shows allele frequencies of the same mutations in Zurich WWTP samples over time, while the right panels show allele frequencies measured in individual biofilm samples. Mutations are ordered by average frequency in the Zurich WWTP samples, and color indicates allele frequency (0-1).

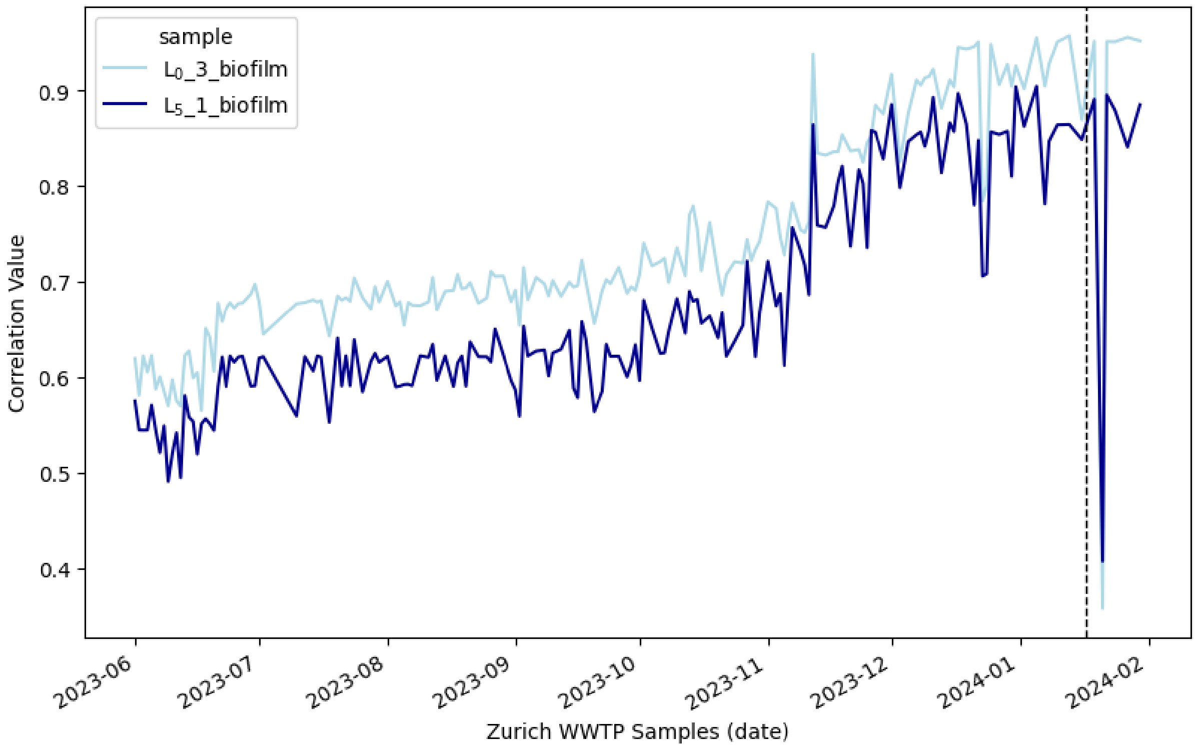


**Figure S6. Correlation of allele frequency profiles of biofilm samples and Zurich WWTP wastewater samples.** The horizontal axis indicates the collection date of wastewater samples from the Zurich WWTP, while the vertical axis shows the Pearson correlation coefficient. The vertical black dashed line marks the biofilm sampling date (i.e., 17 January 2024).

### Supplementary Text

#### Text S1. Estimation of percentage change between sampling locations.

For each target and sampling experiment, we fitted a dPCR-specific generalized linear model with predictor

$$log\ (E[concentration_i])\ = \alpha + \beta \times location(i)$$

to the measured concentrations, where $i$ is the index of each observation. We here modeled the biological replicates as separate observations, using the arithmetic mean of the technical duplicates for each biological replicate. The categorical predictor $location(i)$ models a multiplicative change in the expected concentration for observation $i$ . The dPCR-specific likelihood accounts for the number of valid partitions in the dPCR run (average of 23,639 partitions) and the partition volume ($1.73 \times 10^{-5}$ µL). The GLM was fitted using Markov Chain Monte Carlo via the R package "dPCRfit", based on the stan No-U-Turn-Sampler sampler with 4 chains and 1000 warmup and 1000 sampling iterations each[1]. We translated the posterior samples of the regression coefficients into percentage changes and computed the posterior median and 95% credible intervals.

**Supplementary References**

1 *Stan Development Team. Stan modeling language users guide and reference manual, Version 2.31*, <https://mc-stan.org/docs/2_31/reference-manual/> (2022). Accessed: 2025-08-04.
